# Mixed-method evaluation of an expedited partner therapy take-home medication program: a pilot ED intervention to improve sexual health equity

**DOI:** 10.1101/2023.04.12.23288504

**Authors:** Emily E. Ager, William Sturdavant, Zoe Curry, Fahmida Ahmed, Melissa DeJonckheere, Andrew Gutting, Roland C. Merchant, Keith E. Kocher, Rachel E. Solnick

## Abstract

**Background:** Treatment for partners of patients diagnosed with sexually transmitted infections (STIs), referred to as expedited partner therapy (EPT), is infrequently used in emergency departments (EDs). This was a pilot program to initiate and evaluate EPT through medication-in-hand (“take-home”) kits or paper prescriptions. This study aimed to assess the frequency of EPT prescribing, the efficacy of a randomized best practice advisory (BPA) on the uptake of EPT, perceptions of ED clinicians regarding the EPT pilot, and factors associated with EPT prescribing.

**Methods:** This pilot study was conducted at an academic ED in the midwestern US between August and October 2021. The primary outcome was EPT prescription uptake and the BPA impact was measured via chart abstraction and analyzed through summary statistics and Fisher’s exact test. The secondary outcome of barriers and facilitators to program implementation was analyzed through ED staff interviews (physicians, physician assistants, and nurses). A rapid qualitative assessment method for the analysis of the interviews was employed.

**Results:** Fifty-two ED patients were treated for chlamydia/gonorrhea during the study period. EPT was offered to 25% (95% CI 15%-39%) of patients. EPT was prescribed twice as often (85% vs. 38%; *p*<0.01) when the interruptive pop-up alert BPA was shown. Barriers identified in the interviews included workflow constraints and knowledge of EPT availability. The BPA was viewed positively by the majority of participants.

**Conclusions:** In this pilot EPT program, EPT was provided to 25% of ED patients who appeared eligible to receive it. The interruptive pop-up alert BPA significantly increased EPT prescribing. Barriers identified to EPT prescribing can be the subject of future interventions to improve ED EPT provision.

## INTRODUCTION

In 2020, there were 677,769 cases of gonorrhea, an increase of 111% since 2009,^1^ and 1.58 million cases of chlamydia in the United States (US). Emergency department (ED) visits for bacterial sexually transmitted infections (STIs) in the US are also increasing in frequency.^2^ The ED is a critical access point for STI care. Patients presenting to the ED for a possible STI are more likely to be positive for an STI than those visiting an outpatient clinic.^3^ Treatment of a patient’s partner is also crucial, particularly for female reproductive health, as there is an estimated 14% rate of chlamydia reinfection^4^ which can lead to severe complications.^5^

Expedited partner therapy (EPT) is one method to reduce STI re-infection. EPT is a safe and effective harm-reduction practice of treating the sex partner(s) of patients with STIs without a clinical examination.^6^ EPT is superior in preventing reinfection compared to standard partner referral: previous large multi-site randomized control trials^7,8^ and meta-analyses^9,10^ have found that patients offered EPT had a reduction in persistent or recurrent gonorrhea or chlamydia infections; notably, two of these studies involved ED patients.^7,8^ Additionally, EPT may decrease population increases in chlamydia at a state-level.^11^ EPT is supported by major health organizations^5^ including the American College of Emergency Physicians.^12^ There have been no adverse drug events reported in prior studies of EPT^9^ nor over a decade of monitoring from the California Department of Health.^13^ EPT is now used in most US publicly funded family planning clinics,^14^ but infrequently provided in the inpatient setting^15–17^ or in EDs.^18^ ED medical directors have reported poor knowledge of how to prescribe EPT^18^ and ED clinicians’ ability to prescribe EPT medications can vary greatly.^18^ Many state regulations prohibit EPT medication costs from being charged to an index patient’s health insurance policy. To address the barrier of the partner’s access to STI treatment, two promising approaches for EDs are either to distribute medication-in-hand (“take-home”) kits or paper prescriptions for the patient to give to their sex partner(s). However, research evaluating these approaches in the ED is lacking.

To investigate possible solutions to EPT implementation in EDs, a pilot program at a single ED to dispense both take-home kits and paper prescriptions was evaluated. ED clinicians were interviewed about their perceptions of the pilot and assessed the frequency of EPT prescribing. In addition, the efficacy of a best practice advisory (BPA) to encourage prescribing was examined. Lastly, variations in EPT prescribing by patient demographic factors, health insurance status, clinician type, and STI testing results were explored.

## METHODS

### Setting and Participants

The pilot study was conducted between August and October 2021 at an academic ED in the midwestern US with a patient volume of over 100,000 visits per year. In Michigan, EPT is legal and encouraged by the Michigan Department of Health and Human Services (MDHHS) for chlamydia, gonorrhea, and trichomoniasis.^19^ Before the onset of this pilot study, the MDHHS began a Centers for Disease Control and Prevention (CDC) grant-funded initiative to increase state-wide use of EPT, which included donating EPT medications to several EDs in the state for index patients to deliver to their partner(s) via take-home kits. The EPT medications were based on CDC guidelines on the presumptive treatment of gonorrhea and chlamydia using an oral-only regimen.^6^ The MDHHS received these medications from a pharmacy distributor and repackaged them into pre-labeled kits with information and instructions for EPT. To abide by drug safety regulations regarding the transfer of medications, a T3 document, in which a drug manufacturer details all product information to a new recipient from the drug manufacturer was obtained and approved by the ED pharmacy. The take-home kits were then delivered to the ED in packages for either potentially pregnant (containing cefixime and azithromycin) or not pregnant (containing cefixime and doxycycline) patients. Pregnancy status was based on ED serum or urine testing. This study was approved by the study site’s Institutional Review Board (HUM00199376) and (HUM00196451).

### Pilot EPT program

The mechanism of the EPT pilot is detailed via a swim lane process map (**Figure 1**). As part of a larger quality improvement initiative, a sexual health electronic health record (EHR) orderset was created to assist ED clinicians with ordering laboratory tests and treatment for patients being evaluated for STIs **(Appendix 1)**. The orderset provided a link to an EPT protocol and the following: 1) Standardized EPT prescriptions for printing on plain paper; 2) Progress note to indicate the provision of EPT; and 3) EPT discharge instructions and resources for local low to no-cost sexual health clinics.

**Figure 1.**
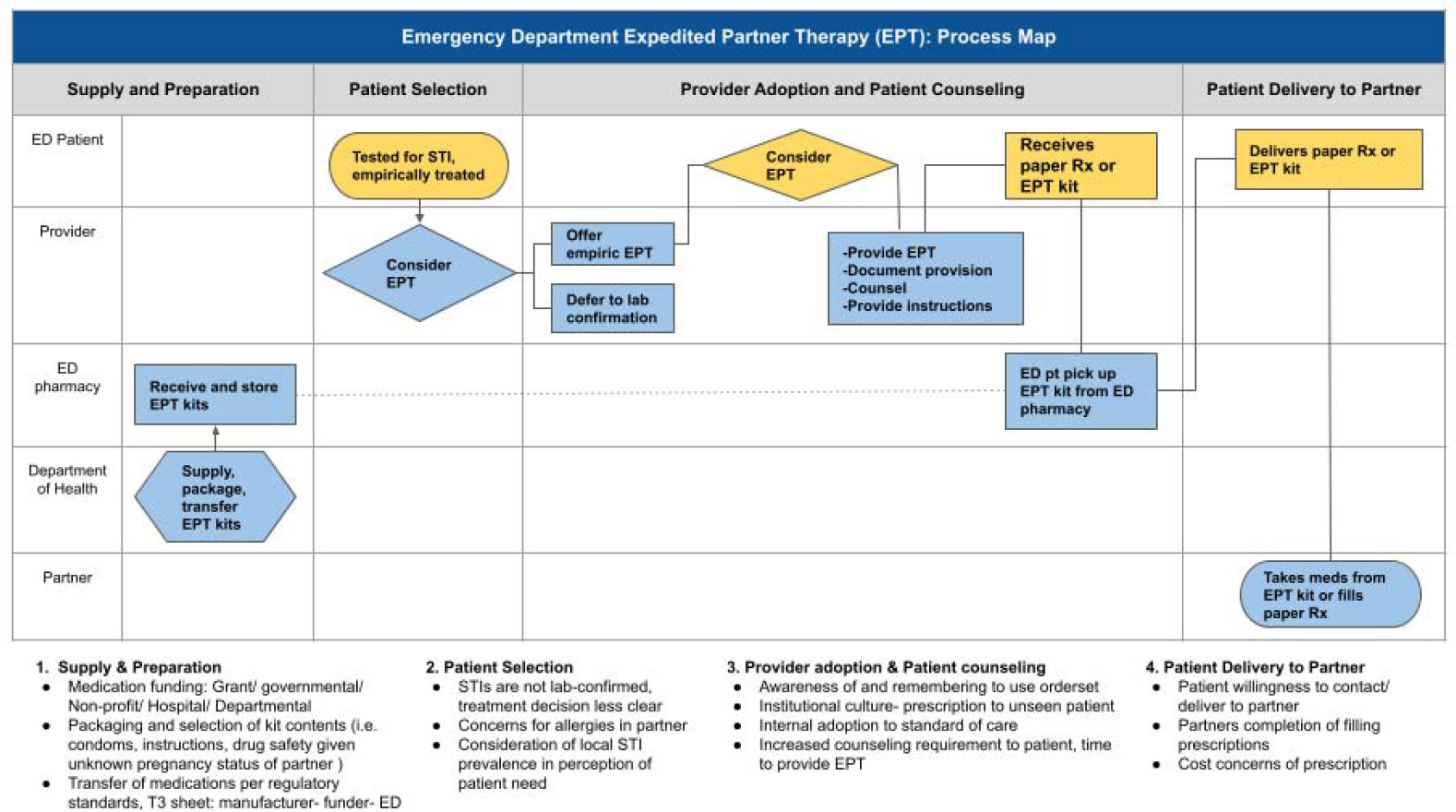
Swim-lane process map of the expedited partner therapy (EPT) pilot program. This swim-lane process map depicts the EPT program responsibilities represented by horizontal swim lanes organized by the role of the stakeholder: ED patient, Clinician, ED pharmacy, Department of Health, and Partner. Based on the interviews and additional discussion with the MDHHS and ED pharmacy over the course of the project, four phases for pilot implementation are displayed across the top of the process map by major activity domains: 1) Supply and Preparation, 2) Patient Selection, 3) Provider Adoption and Patient Counseling, 4) Patient Delivery to Partner. Captions underneath the map summarize key points of the corresponding section.

With assistance from ED pharmacy leadership, an ED-specific protocol was designed to dispense EPT kits. For the “take-home” medication kits, the ED clinician printed the EPT prescriptions, brought the prescriptions to the ED pharmacy located adjacent to the clinician’s workspace, and then the pharmacist took the kit to the patient and provided medication counseling. ED clinicians could offer patients EPT paper prescriptions as an alternative to the kits. At the time of the pilot, Michigan law still allowed plain paper prescriptions for non-controlled substances.^20^ Clinicians could choose either approach per the protocol.

A Best Practice Advisory (BPA) was also created as an interruptive alert designed to appear when ED clinicians empirically treated patients for gonorrhea and chlamydia **(Appendix 2a)**. The interruptive BPA appeared with the following trigger criteria: 1) patient was 18 years or older; 2) gonorrhea or chlamydia test ordered; and 3) ceftriaxone and azithromycin or ceftriaxone and doxycycline were ordered. Metronidazole ordering was initially included as a BPA trigger criterion but was later removed as it too frequently triggered the BPA for patients treated for bacterial vaginosis. Additionally, a non-interruptive alert appeared in the EHR Discharge Navigator for all trigger criteria patients if EPT had not been ordered to notify clinicians that the patient was eligible for EPT **(Appendix 2b)**. Prescribing clinicians received the BPA; nurses did not receive the BPA. To evaluate the efficacy of the interruptive BPA, it was programmed to appear randomly for approximately half of ED visits that met trigger criteria (1:1 randomization of visits, BPA alert, or no BPA alert appearance).

Before the BPA introduction, the pilot EPT program elements were presented at ED faculty, nursing, and physician assistant (PA) meetings and EM residency didactics, followed by emails of presented materials. In addition, ED pharmacy staff posted the protocol and educational materials on a bulletin board in a high-trafficked area of the ED.

### ED staff interviews about the pilot EPT program

Structured interviews were conducted with ED clinicians caring for EPT-eligible patients to explore barriers and reasons for EPT uptake. Purposive sampling was used to interview the attendings, residents, PAs, and nurses who cared for EPT-eligible patients. Each clinician was invited via email to a telephone interview within 72 hours of the patient’s visit. Participants could participate in only one interview, even if they provided care to multiple EPT-eligible patients.

An independently generated, semi-structured interview guide was prepared using elements from the Consolidated Framework for Implementation Research (CFIR)^21^ and published manuscripts on EPT implementation.^22–24^ There is currently no validated interview guide related to EPT. The interview guide was pilot-tested and iteratively revised with three clinicians unaffiliated with this project by conducting cognitive-based assessments using the ‘think-aloud’^25^ approach to ensure comprehension and fidelity to the question intent **(Appendix 3)**. Each telephone interview began with questions on the participant’s background, as well as their EPT knowledge and beliefs. Participants were then asked about their recent EPT-eligible patient encounter, including reasons for or for not prescribing EPT and any barriers or facilitating factors they encountered with the EPT process. The final portion of the interview included questions regarding the BPA. Three multiple-choice questions were also incorporated to introduce each interview topic. Interviews were recorded using a Health Insurance Portability and Accountability Act (HIPAA)-compliant Zoom© audio call, auto-transcribed via Zoom© closed captioning, and saved to a password-protected website for 150 days. Participants were aware of the subject of the interview before agreeing to participate. They were not compensated. Each interview was 10-15 minutes in duration.

The lead researcher was an emergency medicine physician with formal training in qualitative methods and health services research. The interview team was composed of nine individuals: one expert in qualitative methodology who guided the analytical approach (MD), three resident physicians (AK, EA, WS), and four medical students (AR, JL, LD, ZC). All interview team members received training in the rapid assessment qualitative methodology from the lead researcher. The lead researcher did not conduct any of the interviewers. None of the interviewers had a supervisory role related to participants. Interviewers were not compensated.

### Outcomes

The primary outcome was EPT provision and the impact of the BPA on EPT use. Variations in EPT ordering by patient demographic factors, insurance status, clinician type, and STI testing results were examined as an exploratory outcome. The secondary outcomes were barriers and facilitators to EPT program implementation, assessed through ED clinician interviews. These methods are described in further detail below.

### EPT provision analysis

An EPIC (EPIC Systems) Report was created to automatically send same-day daily emails to the research team about ED visits that met the previously described criteria for the interruptive pop-up alert BPA. Each of these ED visits’ EHR Assessment and Plan section was reviewed by a research assistant to confirm that the patient was being treated for a presumed STI instead of another bacterial infection. For ED visits that met these study criteria, research assistants extracted the following data elements: patient demographics (age, gender, race/ethnicity, insurance payer, STI testing results), clinician demographics (resident, attending or PA), and if the clinician was exposed to the interruptive BPA for EPT. Summary statistics are reported with the frequency of each category by EPT ordering. Univariable analysis was conducted with Fisher’s exact test comparing distributions by receipt of EPT. Proportions are calculated with a logit transformed 95% confidence interval. Statistical analyses were performed using STATA (StataCorp, version 16).

### ED staff interviews analysis

Data were analyzed using a rapid assessment method.^26^ During the interviews, researchers paraphrased responses in real-time or transcribed select quotes verbatim immediately following the interview with assistance from the auto-transcription and Zoom© recording. Interviewers also coded the data immediately following each interview. Interviewees were emailed a list of their reported verbatim quotes and asked to comment on the accuracy of their quotes and provide any needed corrections. Codes were created according to a CFIR-based coding scheme and prior relevant EPT literature.^22,23,27,28^ Interviewers iteratively added codes to reflect new ideas not included in the a priori coding scheme until data saturation was achieved. Themes were derived deductively and organized by CFIR domains, with additional themes added based on patterns in the coding elements. Two reviewers (EA, WS) independently evaluated the coded data to identify patterns, while a third (RS) reconciled any discrepancies. The Standards for Quality Improvement Reporting Excellence (SQUIRE 2.0) and Consolidated Criteria for Reporting Qualitative Research (COREQ) reporting guidelines were used as a framework for reporting data (**Appendix 4**).^29,30^

## RESULTS

### EPT provision

During the study period, 52 ED patients were tested and empirically treated for STIs at the study institution. Their demographic characteristics are provided in **Table 1**. Only physician residents or PAs were the prescribing clinician type, which is typical at this academic institution. Of the 52 patients, 14 patients (27%;95% CI, 16%-41%) had a laboratory-confirmed test for either gonorrhea, chlamydia, or trichomoniasis, and 13 patients (25%;95% CI,15%-39%) were provided with EPT. Of the 14 patients with a laboratory-confirmed test for either gonorrhea, chlamydia, or trichomoniasis, three (21%;95% CI, 6%-53%) received EPT. EPT prescribing did not differ by demographics, the type of ED clinician involved in the patient’s ED visit, or whether or not the patient had a laboratory-confirmed test for either gonorrhea, chlamydia, or trichomoniasis. However, EPT was prescribed more than twice as often (85% vs. 38%) when the prescribing ED clinician was randomly shown the interruptive pop-up alert BPA (*p*<0.01).

**Table 1.**
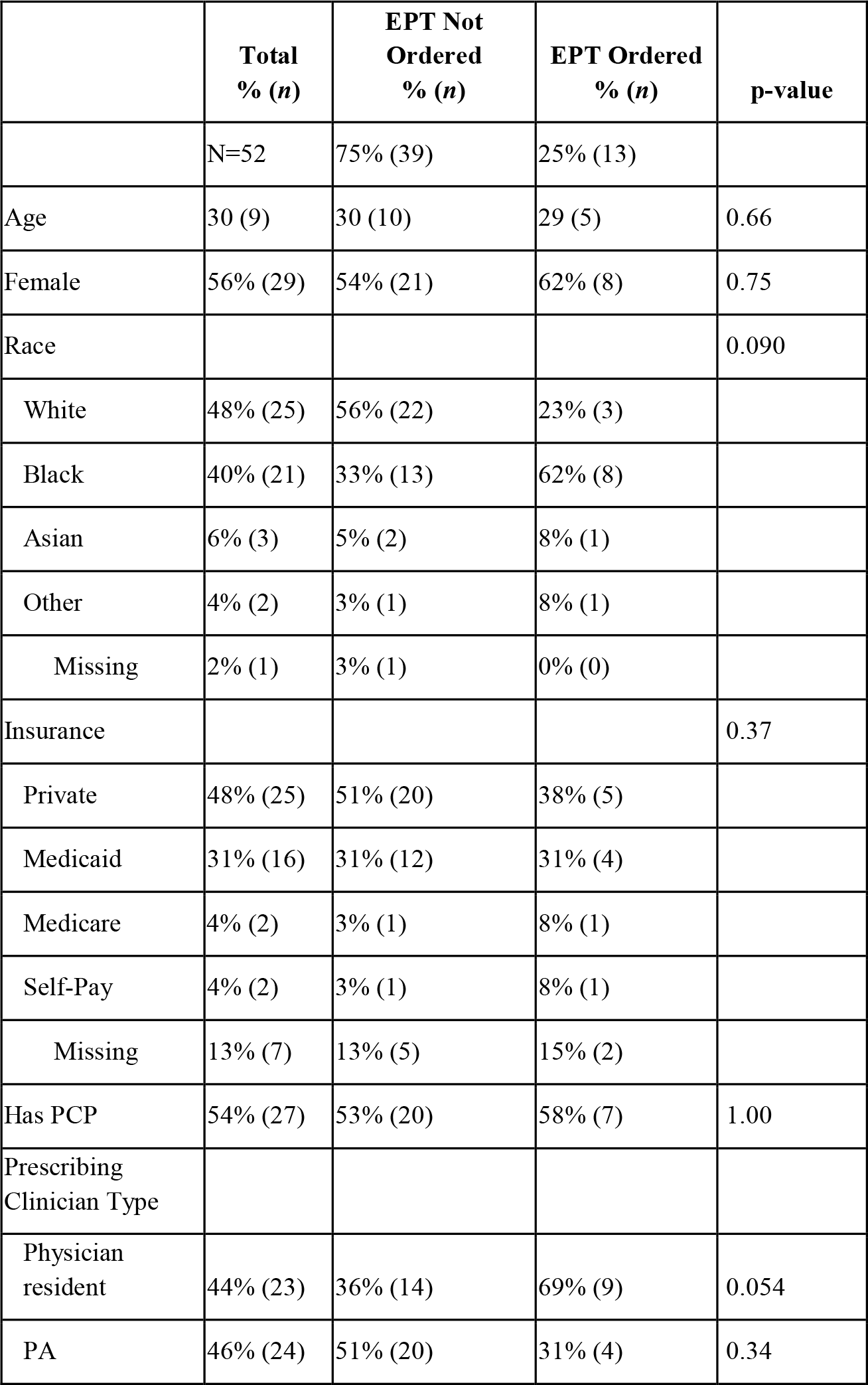
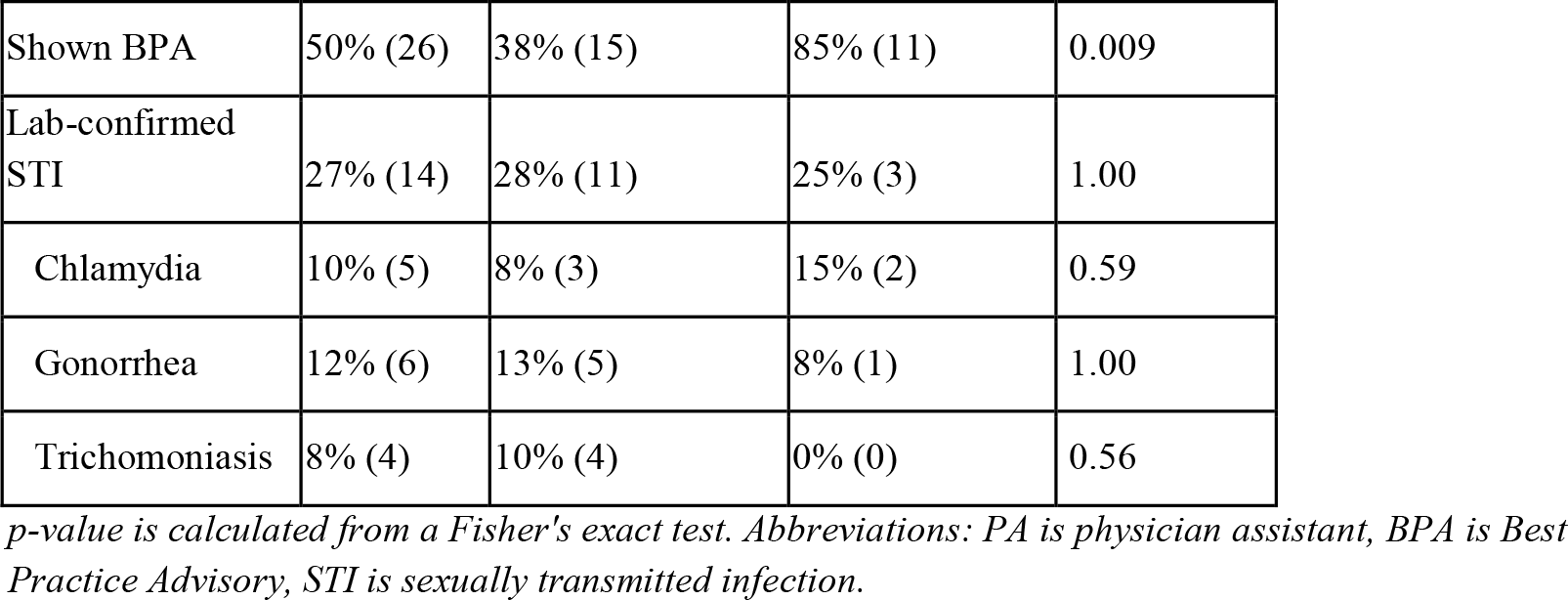
Expedited partner therapy (EPT) utilization and characteristics of ED patients presumptively treated for sexually transmitted infections.

|  | **Total**<br>**% (n)** | **EPT Not**<br>**Ordered**<br>**% (n)** | **EPT Ordered**<br>**% (n)** | **p-value** |
| --- | --- | --- | --- | --- |
|  | N=52 | 75% (39) | 25% (13) |  |
| Age | 30 (9) | 30 (10) | 29 (5) | 0.66 |
| Female | 56% (29) | 54% (21) | 62% (8) | 0.75 |
| Race |  |  |  | 0.090 |
| White | 48% (25) | 56% (22) | 23% (3) |  |
| Black | 40% (21) | 33% (13) | 62% (8) |  |
| Asian | 6% (3) | 5% (2) | 8% (1) |  |
| Other | 4% (2) | 3% (1) | 8% (1) |  |
| Missing | 2% (1) | 3% (1) | 0% (0) |  |
| Insurance |  |  |  | 0.37 |
| Private | 48% (25) | 51% (20) | 38% (5) |  |
| Medicaid | 31% (16) | 31% (12) | 31% (4) |  |
| Medicare | 4% (2) | 3% (1) | 8% (1) |  |
| Self-Pay | 4% (2) | 3% (1) | 8% (1) |  |
| Missing | 13% (7) | 13% (5) | 15% (2) |  |
| Has PCP | 54% (27) | 53% (20) | 58% (7) | 1.00 |
| Prescribing<br>Clinician Type |  |  |  |  |
| Physician<br>resident | 44% (23) | 36% (14) | 69% (9) | 0.054 |
| PA | 46% (24) | 51% (20) | 31% (4) | 0.34 |
| Shown BPA | 50% (26) | 38% (15) | 85% (11) | 0.009 |
| Lab-confirmed STI | 27% (14) | 28% (11) | 25% (3) | 1.00 |
| Chlamydia | 10% (5) | 8% (3) | 15% (2) | 0.59 |
| Gonorrhea | 12% (6) | 13% (5) | 8% (1) | 1.00 |
| Trichomoniasis | 8% (4) | 10% (4) | 0% (0) | 0.56 |

*p-value is calculated from a Fisher's exact test. Abbreviations: PA is physician assistant, BPA is Best Practice Advisory, STI is sexually transmitted infection.*

### ED staff interviews about EPT

Of the 106 ED clinicians invited to be interviewed, 20 participated **(Table 2)**. Of the 20 interviewees: eleven were attending physicians, five were emergency medicine residents, two were PAs, and two were nurses. Thirteen were female. Additional representative quotes are displayed in **Table 3**, and key considerations are summarized by their respective roles on the process map **(Figure 1)**.

**Table 2.** Characteristics of physicians, physician assistants, and nurses who participated in clinician interviews.

| **Participant Characteristic** | **Number (%)** |
| --- | --- |
| Self-identified gender |  |
| Male | 13 (65) |
| Female | 7 (35) |
| Age (years), median (range; IQR) | 35 (27-61; 32-43) |
| 20-29 | 3 (15) |
| 30-39 | 11 (55) |
| 40-49 | 2 (10) |
| 50-59 | 2 (10) |
| 60-69 | 2 (10) |
| 70+ | 0 (0) |
| Role in ED |  |
| MD/MBBS | 16 (80) |
| Physician (attending) | 11 (69) |
| Physician (resident) | 5 (31) |
| Physician assistant | 2 (10) |
| Registered nurse | 2 (10) |
| Years in Practice Post-Residency (years), median (range; IQR) | 3.5 (0-37; 0-13) |
| 0 | 5 (25) |
| 1-9 | 9 (45) |
| 10-19 | 3 (15) |
| 20+ | 3 (15) |

**Table 3.** Representative clinician quotes regarding expedited partner therapy (EPT) organized by CFIR Domain and Constructs.

| CFIR Domain | CFIR Constructs | Topic | Representative quote |
| --- | --- | --- | --- |
| Outer Setting | Patient Needs & Resources, External Policy & Incentives | Improving STI Treatment | *It is super important [to prevent STI reinfection] to public health and to the health of our patients, and to lowering the barriers they have to medical care. (Participant 5)* |
|  |  |  | *[We] see patients who don't interact with the medical system frequently - we are a point of contact for them and have the opportunity to give them this intervention that improves public health. (Participant 20)* |
|  |  |  | *I think it's capturing a group of patients we don't always see in the ED. (Participant 18)* |
|  |  |  | *I think it would be helpful from a community health perspective, and [it] would help the spread of disease. (Participant 10)* |
| Inner setting | Culture, Structural characteristics, Access to Knowledge & Information, Implementation Climate | Unfamiliarity, Increased Workload | *[A barrier is] thinking about doing it. (Participant 5)* |
|  |  |  | *Unfortunately, [a barrier is] just the extra work involved. (Participant 16)* |
|  |  |  | *I still think it's weird to write a prescription for someone I haven't seen as a patient but I am not opposed to doing it. (Participant 1)* |
|  |  |  | *It's not entirely up to the ED but we should play a role and take advantage of the tools available to us, including EPT. (Participant 16)* |
|  |  |  | *The long-term answer is that it becomes [the] standard of care. (Participant 4)* |
| Characteristics of Individuals | Knowledge & Beliefs About the Intervention | Unseen Partner, Uncertain Delivery | *I'm concerned mostly for safety - if they are allergic, [have a] drug reaction, or some type of kidney disease that's undiagnosed. (Participant 15)* |
|  |  |  | *I think if they leave with medications in hand that's better because there's less steps. (Participant 17)* |
|  |  |  | *More options are better. I like the idea of giving medication more as getting medications seems easier. (Participant 5)* |
|  |  |  | *It would be great if it makes it to the intended* individual. Do they actually get the medication? (Participant 12) |
|  |  |  | *What would happen if you gave a script to someone whose allergies you can not check? (Participant 19)* |
| Intervention Characteristics | Complexity, Cost | Take-home Program, Distribution | *This BPA should exist because it is for [an] intervention that, without a hard-stop reminder, it would not be prescribed otherwise. (Participant 20)* |
|  |  |  | *[The] take-home med kit is preferable; [it] reduces extra step for [the] patient to give [the] partner a prescription [and] to fill that prescription. (Participant 20)* |
|  |  |  | *The patient was discharged and was in a hallway spot waiting for 30-40 minutes waiting for the medications; the patient almost left without medications because he was ready to go. (Participant 15)* |
|  | Design Quality & Packaging | Orderset, Best Practice Advisory | *As I recall the orderset was pretty straightforward. (Participant 12)* |
|  |  |  | *BPA's would be the best reminder; if you want people to pay attention and do something you need a BPA. (Participant 8)* |
|  |  |  | *I'm sure it's helpful but more BPA's are painful. The more times I'm interrupted the more I'm likely to make a mistake on the thing I wanted to do. (Participant 4)* |

### Outer Setting (Patient Needs & Resources, External Policy & Incentives)

#### Improving STI Treatment

Many participants noted the public health benefit of EPT and the unique role the ED has in caring for underserved patients. Most remarked that it was important for EDs, in general, to prevent STI reinfection and that EPT is effective in preventing STI reinfection in ED patients.

> *I think this could be really helpful to lots of EDs; we see limited encounters for this indication, but for those we do see it has the potential to be very valuable especially if we bring this to patients in a non-judgmental way. [EPT is] very important - we see lots of folks that don’t have a primary care doctor or gynecologist. We have the opportunity to educate, treat, [and] prevent the long-term sequelae of these types of infections. (Participant 9)*

### Inner setting (Culture, Structural characteristics, Access to Knowledge & Information, Implementation Climate)

#### Unfamiliarity, Increased Workload

Frequently mentioned inner setting characteristics included knowledge of the basic concept of EPT and time constraints in using it. Almost all participants were able to accurately or at least partially define EPT. Though most participants were familiar with the concept, only about half knew how to order EPT at the study site.

> *I didn’t know the program existed and did not know how to use the order set or that it was available. (Participant 8)*

While the majority were supportive of having EPT available as an option, the extra work involved in education was cited by several participants as a barrier. They stated that some patients may not understand STIs and thus need extra counseling. Especially in a busy ED, the extra time involved in bridging these knowledge gaps was undesirable.

> *[A barrier is] explaining to the patient how to explain to partner, which can be challenging…not insurmountable. (Participant 19)*

Culture at the study site was also frequently cited as a barrier. These clinicians stated that EPT is not a common practice and it was challenging to remember to use. Further, several participants viewed EPT as less impactful in EDs with a lower volume of STI visits.

> *It feels strange to write a prescription without a name on it. Giving it to someone you’ve never met or interacted with feels strange; it’s a change in practice. (Participant 1)*

Participants also mentioned ways to change this culture. Two clinicians stated that incorporating EPT into the standard of practice is the best long-term solution for clinicians to order EPT consistently.

> *I think everyone just needs to do it once, and then it will be a part of our practice. The volume of us seeing an exposure needs to grow and then just knowing to do it from now on. (Participant 6)*

### Characteristics of Individuals (Knowledge & Beliefs About the Intervention)

#### Unseen partner, uncertain delivery

One clinician expressed reluctance related to treating a patient’s partner when the index patient had an unconfirmed STI status.

> *A limitation is that we don’t have confirmed test results and so without results, I may at times feel reluctant to send a partner home with a kit. (Participant 5)*

Medication cost when filling paper prescriptions was also listed as a reason for a reluctance to order EPT.

> *I have concerns that the medications cost a lot of money and we may be adding a burden to the patient. (Participant 8)*

A minority of clinicians were concerned about prescribing for someone they hadn’t evaluated as a patient and adverse medication effects or allergies. When asked about the EPT take-home kit, a participant stated,

> *Some [doctors] may be reluctant to do that because they haven’t been able to examine the patient’s partner. (Participant 10)*

### Intervention Characteristics (Complexity, Cost)

#### Take-home Program, Distribution

Participants preferred the take-home EPT kits over paper prescriptions due to the fewer steps for treatment of sex partners but recognized it was likely a shared decision between patient and clinician.

> *It depends on the patient’s situation. You need to discuss with the patient if they are willing or want to dispense medication to a partner or if a prescription will be effective. (Participant 16)*

A process barrier observed by some participants was a delay in the ED pharmacist filling the take-home EPT kit, with one clinician noting that the patient had to wait 30 minutes after discharge for the kit.

Factors outside the control of ED clinicians, including the patient’s actions, were frequently cited as barriers to the implementation of EPT. Half of the participants were concerned that the success of EPT depended on patient factors, such as delivering the take-home kit to their sex partner or filling the paper prescription. They worried that the patient would no longer have contact with their sex partner or would not deliver the take-home kit or prescription to their sex partner.

> *The biggest [barrier] is if they can’t get in touch with their partner again - I don’t even know this partner so I’m not going to give them the prescription. (Participant 6)*

### Intervention Characteristics (Design Quality & Packaging)

#### Orderset, Best Practice Advisory

Participants noted the simplicity of finding and navigating the EPT EHR orderset. When asked “what went well with the EPT process in the ED,” a quarter indicated it was “easy to use.” Participants stated that the orderset was straightforward, well-designed, and easy to find in the EHR. The EPT-specific discharge instructions were also considered efficient and helped with patient education.

Perspectives on BPAs were mixed but generally positive. Almost half of the participants suggested using a BPA when asked how to implement ED-based EPT. When asked, “how do you feel about a reminder BPA for EPT that pops up when you order empiric therapy,” there were thirteen responses for “appreciate it as a reminder,” seven responses for “like,” and four responses for “don’t like.” Participants also stated that they support BPAs that are more “patient-centered” rather than intended for financial or medico-legal purposes.

> *I think the BPA is the most streamlined way to do it; if there wasn’t a hard stop I may not have written the prescription in this circumstance; I know some people are against BPAs in general but I liked it. I think if we’re really trying to implement this a hard-stop BPA is the best way to not miss these prescriptions. It is probably the most effective way. (Participant 18)*

Several participants had conflicted feelings towards a “hard-stop” BPA, stating that clinicians already encounter numerous BPAs which can be disruptive to clinician workflow. Conversely, several stated they would support a “hard-stop” BPA since the intervention may be otherwise forgotten.

> *I’m conflicted about it. I dislike BPAs probably because of how many we have. So adding another one makes me cringe. But if there’s a way to do it at the time of discharge rather than during the encounter then I wouldn’t hate it. (Participant 17)*

Additional non-BPA-based suggestions for early adoption included: encouraging attendings to remind residents to use EPT, more heavily involving the ED pharmacist in the EPT process, and having a “standing nursing order” to order EPT medications.

## DISCUSSION

This pilot study demonstrates the feasibility of a novel ED-based EPT take-home medication pilot program and the effectiveness of EHR interventions to facilitate adoption. EPT use among all patients being presumptively treated for gonorrhea or chlamydia was examined, as well as the efficacy of an interruptive BPA for EPT. While the BPA greatly increased EPT prescribing, EPT was only offered to 25% of EPT-eligible ED patients. This low level of EPT prescribing was surprising, especially given that most interviewees accurately conveyed the concept of EPT and supported its provision from the ED.

The low level of EPT ordering may be due to several factors. First, there was an educational gap in how to order EPT: half of the participants stated that they did not know how to order EPT. Second, patients offered EPT may have declined, which this study could not measure. In a survey of pediatric ED patients, participants uninterested in EPT cited that they were concerned for partner safety, wanted the partner to get a diagnosis, or felt EPT would detract from the partner’s accountability.^31^ Third, low use may be related to logistical difficulties involved in providing take-home medications. Though clinicians preferred take-home kits over paper prescriptions, they also recalled the long process of kit distribution, possibly due to pharmacists’ unfamiliarity with the process. In the future, this will be an essential consideration, as delays in providing the medication in hand are a potential back-end issue. Addressing this concern may not only increase order rates among providers but increase efficiency and prevent delays in care. Fourth, ED clinicians could have forgotten to prescribe EPT, as evidenced by interviewees who reported that remembering to prescribe EPT was challenging. The interruptive BPA likely reduced this issue. The non-interruptive BPA in the Discharge Navigator used in this project was probably overlooked, as none mentioned seeing it.

Despite mixed feelings among ED clinicians towards BPAs in general, the majority supported an EPT-specific BPA, with nearly half suggesting using a BPA to increase EPT ordering. Further, the over two-fold increase in EPT ordering during BPA-exposed visits supports its efficacy for EPT implementation. This study adds to the growing research on clinician acceptability^32^ of using BPAs to improve patient care.^33–37,38^ A study from an urban ED including 75,901 patients demonstrated that a targeted BPA increased syphilis screening by 124% compared to clinician-initiated testing.^39^ However, BPAs are known to contribute to alarm fatigue^40^ and must be designed to minimize inappropriate interruptions.^41^ Future work on ED EPT may investigate when to discontinue a BPA as clinician familiarity increases.

Only three patients out of the 13 provided EPT had a lab-confirmed STI, suggesting presumptive EPT may lead to overprescribing. A potential solution to improving the accuracy of ED-based EPT provision is rapid testing for chlamydia and gonorrhea.^42–45^ On the other hand, EPT could still be appropriate even when laboratory testing is negative if the patient was tested before the test could accurately detect the infection (i.e., a “window period” after exposure). Without rapid testing, many EDs have developed dedicated follow-up teams to address positive test results after ED discharge. Offering EPT during such follow-up interactions could help target EPT to patients with lab-confirmed infections.

An innovative aspect of this EPT program was the ability to provide “take-home” EPT kits. While EPT has been demonstrated to increase follow-up rates with the index patient’s partner and reduce reinfection rates among the index patient,^46^ the total rate of treatment of the patient’s partner is still relatively low, owing in part due to low prescription filling rates, sometimes found to be less than 50%.^47,48^ This study demonstrates one method for addressing this back-end issue: providing take-home medications rather than a written prescription. This would reduce the impact of one major limiting factor in the completion of treatment. For this reason, the CDC recommendations on EPT state a preference for take-home medications.^6^ Other studies have shown the benefit of “med to bed” or “take-home” programs to facilitate medication compliance when there is concern about a patient’s access, such as in anticoagulants and medications for opioid use disorder.^49–51^ Though other ED “take-home” medications may be charged to a patient’s healthcare insurance plan, this mechanism is not currently possible for EPT, as most health insurers will not pay for medication for anyone other than the covered individual. One known exception is California, where, since February 2020, the state Medicaid provider – including

Medi-Cal and Family PACT insurance – must cover partner EPT medications for low-income patients.^52,53^ Additionally, certain family planning clinics, health department STI clinics, and Federally Qualified Health Centers pay for EPT medications via governmental grants.^54^ Given disparities in healthcare access among patients who receive STI care in EDs,^55,56^ expanding ED-EPT medication funding and California’s Medicaid regulations to other states would increase EPT provision.

## LIMITATIONS

Limitations of this study include the small sample size, which reduced the ability to identify differences between groups if they existed. External validity is limited because the study was conducted at one ED whose population, setting, and resources may be different from other settings. The overall low response rate to the interview invitation also introduces the potential for selection bias. Due to limitations in data collection based on daily EHR reports, data were collected only on how many patients received EPT rather than how many patients may have been offered but then declined EPT. In addition, the ability of this EPT program to provide paper prescriptions could be unique. Electronic medication prescription is the norm in the US,^57^ although some states allow paper prescriptions for EPT.^58^ In addition, the BPA did not trigger for patients being treated for trichomonas, as this infection was too infrequently diagnosed in the ED and keeping metronidazole as a trigger criterion significantly decreased the sensitivity of the screening for eligible patients.

Given the limits of this pilot study methodology, patients’ compliance to provide the “take-home” kits or paper prescriptions to their partners, if the paper prescriptions were filled, or if the medications were taken is unknown. The lack of follow-up limited the ability of this study to confirm medication adherence and completion of treatment in this patient population. Though prior assessments of follow-up and completion of treatment have corresponded to a number needed to treat of 34^7^, this study lacks the follow-up to make any conclusions on NNT or completion of treatment once provided.

## CONCLUSION

In summary, in this pilot EPT program, EPT was provided to 25% of ED patients who appeared eligible to receive it. The interruptive BPA increased EPT prescribing more than two-fold. Multiple barriers to EPT prescribing from this ED were identified, which can be the subject of future interventions to improve ED EPT provision.

## Data Availability

All data produced in the present study are available upon reasonable request to the authors.

