## Supplemental Figure 1 for "Mixed-method evaluation of an expedited partner therapy take-home medication program: a pilot ED intervention to improve sexual health equity"

*Appendix 1.* *Expedited partner therapy (EPT) order set and paper prescription.*

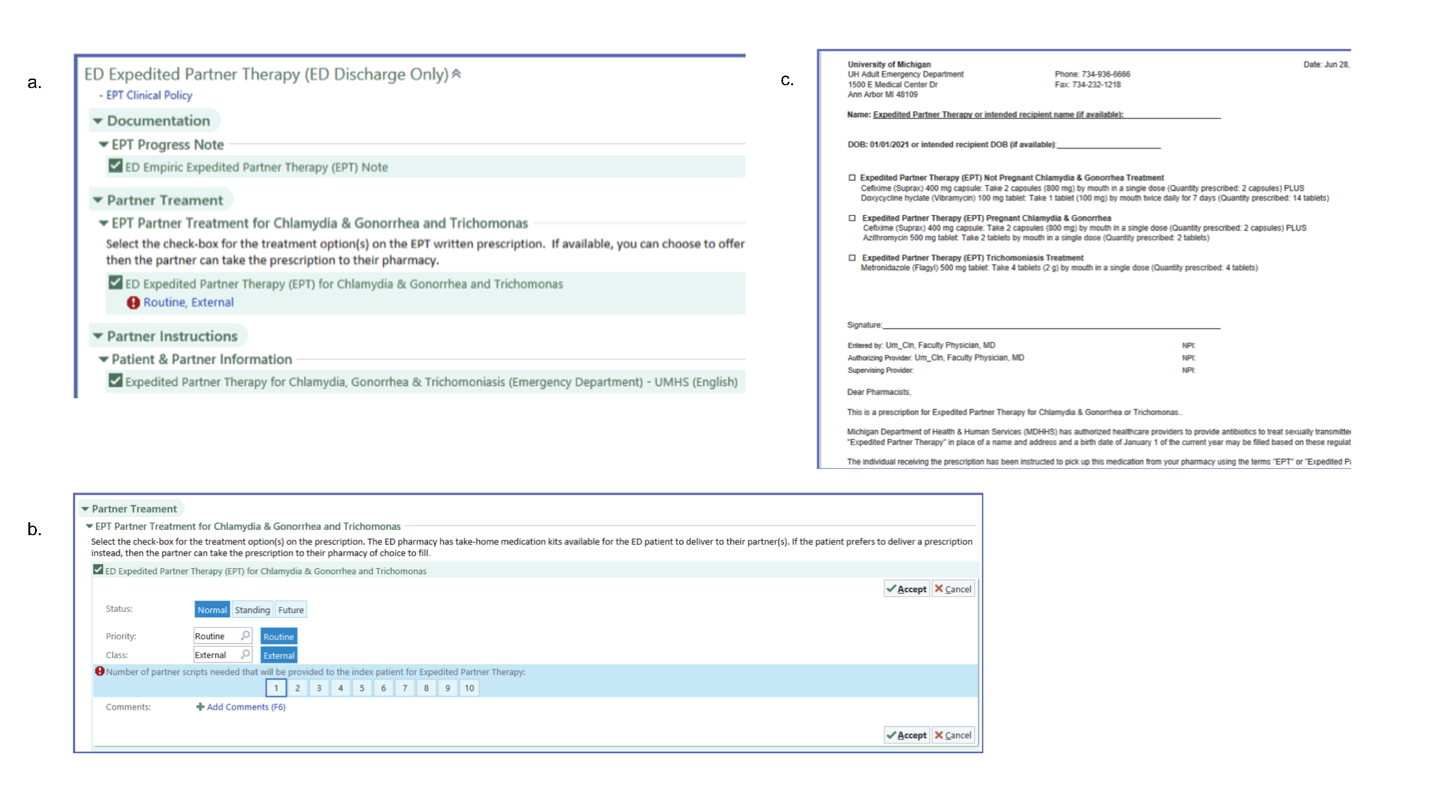

*EPT resources implemented to assist EPT ordering during ED patient encounters, including* ***a)*** *EPT EHR orderset with a link to new EPT protocol,* ***b)*** *standardized EPT prescriptions, and* ***c)*** *EPT paper prescription print-out.*
