## Supplemental Figure 2 for "Mixed-method evaluation of an expedited partner therapy take-home medication program: a pilot ED intervention to improve sexual health equity"

*Appendix 2.* *Expedited partner therapy (EPT) best practice advisory (BPA).*

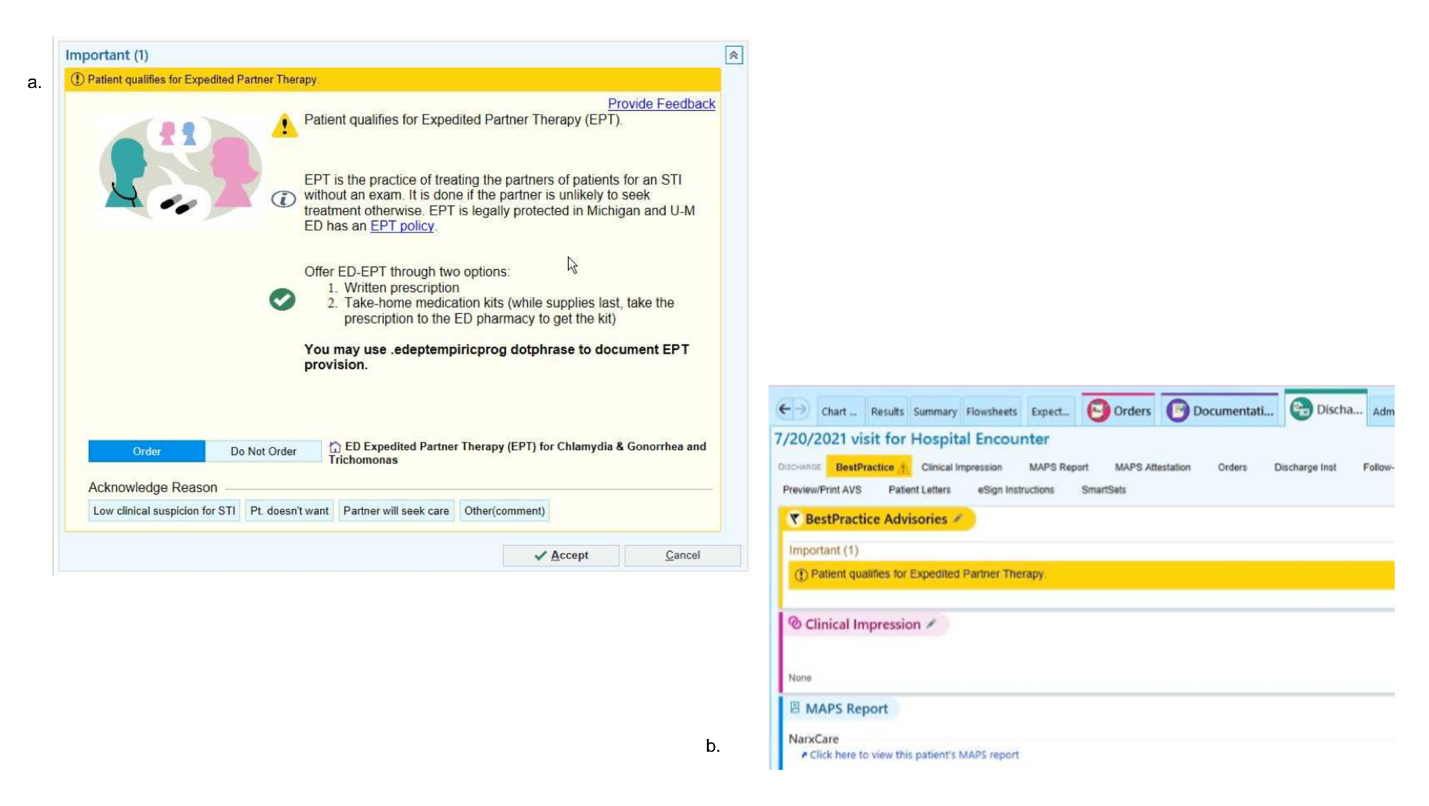
*EPT resources implemented to prompt EPT ordering by ED clinicians, including* ***a)*** *interruptive BPA for EPT and* ***b)*** *non-interruptive BPA for EPT in Electronic Health Record (EHR) discharge navigator.*
