## Supplemental File 3 for "Mixed-method evaluation of an expedited partner therapy take-home medication program: a pilot ED intervention to improve sexual health equity"

### Intro

*Q1.* Interviewer: Please select your name from this list

*Q2.* If previous selection was "Other", please type your name as the interviewer:

*Q3.* Thank you so much for consenting to take this survey about STI care in the ED. We hope to learn more about provider perspectives on partner therapy. Did you have any questions on the written consent we emailed you? As a reminder, the information that I receive from will be strictly confidential. You may skip any question you do not wish to answer.

We would like to audio record the interview so I can take more accurate notes. Once the study is complete all audio recordings will be deleted. May I also have your consent for the recording? **[If no, they may still participate "You may still participate without the recording."]**

[Note to interviewer= when you finish survey questions, do not submit until you use back arrows and complete the post survey codes]

[Note to interviewer- insert the interviewees name here, dont ask them for it]

______________________________

**Background**

*Q4.* "First we have some questions about your background"

What is your degree?

- ○ MD
- ○ DO
- ○ PA
- ○ RN
- ○ ______________ other

*Q5.* How old are you?

______________________________

*Q6.*
How many years have you been in practice post residency? [Can give approximate]

______________________________

*Q7.*
What is your gender? [open-ended]

- ○ Male
- ○ Female
- ○ Non-binary / third gender
- ○ Prefer not to say

### EPT and visit

*Q8.* "Now I have a few questions about EPT in general."

Can you tell me your understanding of the definition of EPT (expedited partner therapy)?

*[If they answer incorrectly, please correct them: "Expedited Partner Therapy (EPT) is the clinical practice of treating the sexual partners of patients diagnosed with chlamydia/ gonorrhea/ or trichomonas) by providing prescriptions or medications to the patient to take to his/ her partner(s) without a healthcare provider first examining the partner. In Michigan it is recommended by for either lab-confirmed or suspected STIs.*

*Use the multiple choice to mark if they were correct or not*

- ○ Correct
- ○ Incorrect
- ○ Partial correct
- ○ [ ] Other

*Q9.* [Interviewer: In what way was the answer not correct? ]

[ ]

*Q10.* "I'm going to ask you about a couple of different EPT interventions."

"At U-M, we just launched a take-home medication kit pilot, where you can print out a prescription for EPT through an order set and then have the ED pharmacist bring your patient a pre-dosed medication kit with directions for their partner."

What are your thoughts on the take-home medication kit program?

Paraphrase or use "Quotes"

Post-survey summary

*Q11.* [Post survey potential codes] *What are your thoughts on the take-home medication kit program?*

- ☐ (barrier) medicolegal - allergy or drug interactions
- ☐ (barrier) medicolegal- worry about getting sued
- ☐ (barrier) medicolegal- worry about missed diagnosis
- ☐ (barrier) "personal responsibility" want partners to get care themselves
- ☐ (barrier) concerned about who funds the medications, sustainable?
- ☐ (barrier) concerned that it will go to wrong person
- ☐ (barrier) concern it will get sold
- ☐ (barrier) worry about partner violence
- ☐ (barrier) takes too much time to talk about/ busy on shift
- ☐ (barrier) diagnostic uncertainty- don't know if pt truly positive for STI
- ☐ (barrier) not ED's place, clinics should do this
- ☐ (barrier) create bad incentive for the pt to return to ED with same problem
- ☐ (benefit) ensures medicine gets to partner
- ☐ (benefit) increase compliance
- ☐ (benefit) reduces spread of infection/ public health mission
- ☐ (benefit) reduces exposure for pt / reduces reinfection
- ☐ (benefit) reduces barrier to get medicine
- ☐ (benefit) addresses issues of affordability of meds
- ☐ (benefit) similar to naloxone
- ☐ (benefit) prevent consequences of untreated STI ( ie PID)
- ☐ (benefit) ED should do more public health

- [ ] preferred to written prescription
- [ ] follow-up nurse should do instead once STI dx confirmed
- [ ] ________ other

*Q12.*
"At the U-M ED, you also have the option to give them a written-prescription and instructions printed out from an order set. This prescription is printed out nameless, without a patient name written, so the patient can deliver it to anyone. Multiple prescriptions can be printed."

What are your thoughts on the written prescription?

Paraphrase or use "Quotes"

Post-survey summary

*Q13.* [Post survey potential codes] **Note these are the same codes/ order as the codes for take-home kit ** *What are your thoughts on the written prescription?*

- [ ] (barrier) medicolegal - allergy or drug interactions
- [ ] (barrier) medicolegal- worry about getting sued
- [ ] (barrier) medicolegal- worry about missed alternative/ diagnosis
- [ ] (barrier) "personal responsibility" want partners to get care themselves
- [ ] (barrier) concerned about who funds the medications, sustainable?
- [ ] (barrier) concerned that it will go to wrong person
- [ ] (barrier) concern it will get sold
- [ ] (barrier) worry about partner violence
- [ ] (barrier) takes too much time to talk about/ busy on shift
- [ ] (barrier) diagnostic uncertainty- don't know if pt truly positive for STI

- ☐ (barrier) not ED's place, clinics should do this
- ☐ (barrier) create bad incentive for the pt to return to ED with same problem
- ☐ (benefit) ensures medicine gets to partner
- ☐ (benefit) increase compliance
- ☐ (benefit) reduces spread of infection/ public health mission
- ☐ (benefit) reduces exposure for pt / reduces reinfection
- ☐ (benefit) reduces barrier to get medicine
- ☐ (benefit) addresses issues of affordability of meds
- ☐ (benefit) similar to naloxone
- ☐ (benefit) prevent consequences of untreated STI ( ie PID)
- ☐ (benefit) ED should do more public health
- ☐ follow-up nurse should do instead once STI dx confirmed
- ☐ preferred to take-home kit
- ☐ ________ other

*Q14.* “Now, we have some questions about your recent U - M ED patient that was empirically treated for an STI”

Can you tell me a little bit about the case and why you empirically treated for an STI?

[If nurse, "why do you think the physician or physician assistant empirically treated for an STI"]

| |
|---|
| Paraphrase or use "Quotes" |
| Post-survey summary |

*Q15.* [Post survey codes] *Can you tell me a little bit about the case and why you empirically treated for an STI?*

- ☐ history was concerning
- ☐ symptoms were concerning
- ☐ exam was concerning
- ☐ worried pt had low healthcare access
- ☐ [ ] other

*Q16.* Do you know how to order EPT at the U-M ED?

*[Interviewer: If they ask, they can order it through the "Discharge" tab using the EPT order smartset ] If it seems like they aren't sure, write how they described it in "Unsure" selection*

- ○ Yes
- ○ No
- ○ [ ] Unsure

*Q17.*
Did you offer EPT?

[If nurse, "was EPT ordered by the MD/ PA"?]

- ○ Yes
- ○ No
- ○ Unsure
- ○ [ ] Other

*Q18.* Why did you offer partner therapy?

Paraphrase or use "Quotes"

Post survey summary

*Q19.* [post survey codes] *Why did you offer partner therapy? *note some of these codes" (benefit) copied from prior code list**

- ☐ high risk patient
- ☐ high risk patient partner
- ☐ low healthcare access
- ☐ just wanted to try EPT
- ☐ no rationale provided
- ☐ ______ other
- ☐ (benefit) ensures medicine gets to partner
- ☐ (benefit) reduces spread of infection/ public health mission
- ☐ (benefit) reduces exposure for pt / reduces reinfection
- ☐ (benefit) reduces barrier to get medicine
- ☐ (benefit) addresses issues of affordability of meds
- ☐ (benefit) prevent consequences of untreated STI ( ie PID)
- ☐ (benefit) increase compliance

*Q20.* **[If applicable]** Why did you NOT offer partner therapy?

*If they say "I forgot", ask "If you had remembered, or been reminded, would you have?"*

Paraphrase or use "Quotes"

Post-survey summary

*Q21.* [if previous response completed]

[post survey codes]*Why did you NOT offer partner therapy?*

- [ ] low risk patient - not confident it was STI
- [ ] concern for HIV/ syphilis in partner
- [ ] concern for other more severe diagnosis in partner not treatable w EPT
- [ ] pt was men who has sex with men
- [ ] forgot
- [ ] couldn't figure out how to order
- [ ] pt said partner was reliable to seek treatent
- [ ] pt won't see partner again
- [ ] pt did not want to
- [ ] i was too busy on shift
- [ ] worried partner was pregnant
- [ ] unaware of data to support
- [ ] other medicolegal concerns
- [ ] other patient safety concerns
- [ ] __________ other

*Q22.* What went well with the EPT processes in the ED?

[If nurse, ask specifically about discharge process if they don't bring it up]

Paraphrase or use "Quotes"

Post-survey summary

*Q23.* [post survey code] *What went well with the EPT processes in the ED?*

- ☐ easy to use
- ☐ pt learned more about sti
- ☐ pt got what they wanted
- ☐ ED team members knew how to do it
- ☐ ED team members wanted it
- ☐ ______ other

*Q24.* What barriers did you encounter with the EPT processes?

If nurse, specifically ask about discharge process.

[Probe: How do you think ED patients responded to being offered EPT? ]
[Probe: How complicated is offering EPT?]
[Probe: What barriers do you think your ED patient will face in participating?]

Paraphrase or use "Quotes"

Post-survey summary

*Q25.* [post survey codes] *What barriers did you encounter with the EPT processes?*

- ☐ almost forgot to order
- ☐ other ED team members did know what EPT was
- ☐ pt had too many questions
- ☐ took too long/ too busy
- ☐ confused on how to order in EMR
- ☐ ________ other

*Q26.* Now I have a few questions about ordering EPT.

Did you see a BPA for EPT for your encounter?

- ○ Yes
- ○ No
- ○ Unsure

*Q27.* How do you feel about a reminder BPA for EPT that pops up when you order empiric therapy?

What about one that is written in the background of the EMR in discharge order set?

*[try for some direct quotes here]*

Paraphrase or use "Quotes"

Post-survey summary

*Q28.* [post survey code] *How do you feel about a reminder BPA for EPT that pops up when you order empiric therapy?*

- ☐ dont like
- ☐ like
- ☐ appreciate it is a reminder
- ☐ it will be ignored
- ☐ ________ other

*Q29.* What would you suggest as a way to be reminded to consider EPT?

**last**

*Q30.* Generally speaking, how do you think EPT will meet the needs of ED patients?

[try to get some direct quotes here]
*If there is not much content, consider these probes- only as a backup:*
*1) Prevent STI reinfection of ED patient /*
*2) Increase access to marginalized population/*
*3) Address untreated STI/*
*4) Prevent sequela of untreated STI such as PID*

Paraphrase or use "Quotes"

Post-survey summary

*Q31.* [post survey code]- *Generally speaking, how do you think EPT will meet the needs of ED patients?*

- ☐ 1) Prevent STI reinfection of ED patient /
- ☐ 2) Increase access to marginalized population/
- ☐ 3) Address untreated STI/
- ☐ 4) Prevent sequela of untreated STI such as PID
- ☐ increase compliance
- ☐ ____________ other

*Q32.* Generally speaking, what are some of the barriers of using EPT for ED patients?

*If there is not much content, consider these probes:*
*1) Patient safety ( ie not actually positive for STI, allergies, missed alternative diagnosis)*
*2) Concern for intimate partner violence*
*3) Legal liability /*
*4) Affordability of EPT medication/*

*5) Ability for pharmacy to fill Rx/*
*6) Unaware of data to support*

Paraphrase or use "Quotes"

Post-survey summary

*Q33.* [post survey code] *Generally speaking, what are some of the barriers of using EPT for ED patients?*

- ☐ what if partner is pregnancy
- ☐ What if they cant afford to fill prescription
- ☐ WHat if pharmacy won't fill prescription
- ☐ What if partner does not want it
- ☐ What if partner is non-compliant/ doesnt complete course
- ☐ Unaware of data to support
- ☐ (barrier) medicolegal - allergy or drug interactions
- ☐ (barrier) medicolegal- worry about getting sued
- ☐ (barrier) medicolegal- worry about missed alternative/ diagnosis ( ie HIV syphilis, other more severe diagnosis like PID)
- ☐ (barrier) "personal responsibility" want partners to get care themselves
- ☐ (barrier) concerned about who funds the medications, sustainable?
- ☐ (barrier) concerned that it will go to wrong person
- ☐ (barrier) concern it will get sold
- ☐ (barrier) worry about partner violence
- ☐ (barrier) takes too much time to talk about/busy on shift
- ☐ (barrier) diagnostic uncertainty- don't know if pt truly positive for STI

- [ ] (barrier) not ED's place, clinics should do this
- [ ] (barrier) create bad incentive for the pt to return to ED with same problem
- [ ] ______ other

*Q34.* To what extent do you agree or disagree with this statement:
**Emergency Medicine has a critical role to play in public health.**

- ( ) (1) Strongly disagree;
- ( ) (2) Disagree;
- ( ) (3) Neither agree nor disagree;
- ( ) (4) Agree;
- ( ) (5) Strongly agree

*Q35.* How important do you think it is for the ED to prevent STI reinfection ?

*Q36.* [post survey code] *How important do you think it is for the ED to prevent STI reinfection ?*

- [ ] not our space
- [ ] should be other healthcare responsibility
- [ ] important
- [ ] not important
- [ ] ______ other

*Q37.*

Do you think that providing **EPT is clinically effective for preventing STI reinfection in ED patients?**

*Q38.* [post survey code] *Do you think that providing EPT is clinically effective for preventing STI reinfection in ED patients?*

- yes
- no
- unsure
- ______ other

*Q39.* Do you think that EDs should be doing EPT?

Paraphrase or use "Quotes"

Post-survey summary

*Q40.* [post survey code] *Do you think that EDs should be doing EPT?*

- yes
- no
- unsure
- ______ yes but only if:

*Q41.* Do you have any additional thoughts on partner therapy?

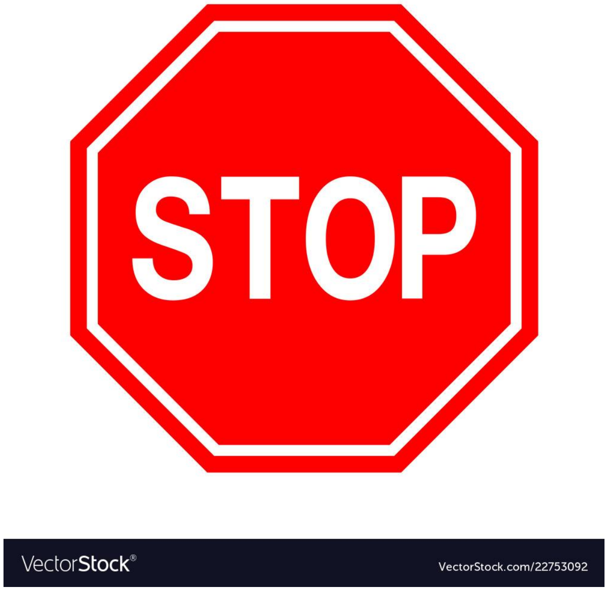

*Q42.*

DO NOT SUBMIT SURVEY OTHERWISE YOU WLL NOT BE ABLE TO GO BACK!!!

PLEASE USE BACK ARROWS NOW TO COMPLETE SUMMARIES AND CODING BEFORE SUBMITTING

*Q43.* What is the MRN for the patient in this encounter?

Powered by Qualtrics
